# Genomic and transcriptomic data analyses highlight *KPNB1* and *MYL4* as novel risk genes for congenital heart disease

**DOI:** 10.1101/2022.01.07.22268881

**Authors:** Martin Broberg, Minna Ampuja, Samuel Jones, Tiina Ojala, Otto Rahkonen, Riikka Kivelä, James Priest, FinnGen, Hanna M. Ollila, Emmi Helle

**Affiliations:** Stem Cells and Metabolism Research Program, Faculty of Medicine, University of Helsinki, Helsinki, Finland; Institute for Molecular Medicine Finland (FIMM), HiLIFE, University of Helsinki, Helsinki, Finland; New Children’s Hospital, Pediatric Research Center, Helsinki University Hospital, Helsinki, Finland; Wihuri Research Institute, Helsinki, Finland; Faculty of Sport and Health Sciences, University of Jyväskylä, Jyväskylä, Finland; Stanford University School of Medicine, Stanford, CA, USA; Center for Genomic Medicine, Massachusetts General Hospital, Boston, MA., USA and Program in Medical and Population Genetics, Broad Institute, Cambridge, MA, USA; Department of Anesthesia, Critical Care and Pain Medicine, Massachusetts General Hospital and Harvard Medical School, Boston, Massachusetts, USA

## Abstract

Congenital heart defects (CHD) are structural defects of the heart affecting approximately 1% of newborns. CHDs exhibit a complex inheritance pattern. While genetic factors are known to play an important role in the development of CHD, relatively few variants have been discovered so far and very few genome-wide association studies (GWAS) have been conducted. We performed a GWAS of general CHD and five CHD subgroups in FinnGen followed by functional fine-mapping through eQTL analysis in the GTEx database, and target validation in human induced pluripotent stem cell - derived cardiomyocytes (hiPS-CM) from CHD patients. We discovered that the *MYL4-KPNB1* locus (rs11570508, beta = 0.24, P = 1.2×10^−11^) was associated with the general CHD group. An additional four variants were significantly associated with the different CHD subgroups. Two of these, rs1342740627 associated with left ventricular outflow tract obstruction defects and rs1293973611 associated with septal defects, were Finnish population enriched. The variant rs11570508 associated with the expression of *MYL4* (normalized expression score (NES) = 0.1, P = 0.0017, in the atrial appendage of the heart) and *KPNB1* (NES = -0.037, P = 0.039, in the left ventricle of the heart). Furthermore, lower expression levels of both genes were observed in human induced pluripotent stem cell derived cardiomyocytes (hiPSC-CM) from CHD patients compared to healthy controls. Together, the results demonstrate *KPNB1* and *MYL4* as in a potential genetic risk loci associated with the development of CHD.

## Introduction

Congenital heart defects (CHD) are globally the most common birth defect, and include a wide range of structural malformations of the heart and great vessels, which have their origin in the first trimester of pregnancy (Huisenga et al., 2021; Liu et al., 2019). CHD may occur as an element of a syndrome or as an isolated defect. Although monogenic cases have been reported, most isolated CHD are thought to be complex traits with reduced penetrance and variable expressivity (Kerstjens-Frederikse et al., 2016). In addition to genetics, environmental exposures such as maternal obesity, diabetes, and certain medications increase the risk for CHD in the offspring (Helle and Priest, 2020; Øyen et al., 2016; Patorno et al., 2017; Persson et al., 2019). Next-generation sequencing studies have identified that the haploinsufficiency of NOTCH*-* and VEGF-signaling pathway genes associate with left ventricular outflow tract obstruction (LVOTO) defects and Tetralogy of Fallot (TOF), respectively (Garg, 2006; Helle et al., 2019, p. 1; Reuter et al., 2019). In most isolated CHD cases, however, the genetic etiology remains to be identified.

As genome-wide sequence data are available for large populations, genome-wide association studies (GWAS) have been used as an additional tool to identify genetic determinants of CHD (Broberg et al., 2021). A number of studies have identified genome-wide significant loci for all CHD (rs185531658)(Lahm et al., 2021) as well as for certain CHD subgroups, such as rs11065987 in Tetralogy of Fallot (Cordell et al., 2013), rs72820264 in conotruncal heart defects and left ventricular obstructive tract defects (Agopian et al., 2017), and rs387906656 in coarctation of the aorta (Bjornsson et al., 2018). As genetic and phenotypic data for hundreds of thousands of individuals has become integrated into large biobank-based projects such as the FinnGen Biobank, UK Biobank or Japan Biobank, the statistical strength for population-wide GWAS has increased (https://finngen.gitbook.io/documentation/).

Here, we conduct a GWAS of 2436 CHD cases from FinnGen release 7 (R7). Our analyses identify one genome-wide significant signal for the total CHD group and a total of four genome-wide significant signals for the following subgroups; early aortic valve disease (aortic stenosis or insufficiency diagnosis at < 60 years), septal defects, and LVOTO defects. Using public tissue-specific expression Quantitative Trait Loci (eQTL) data, we detected significant impact of the CHD associated SNP on the expression of *MYL4* and *KPNB1* in the heart. In addition, we defined the expression of these genes in human induced pluripotent stem cell derived cardiomyocytes (hiPS-CM) from CHD patients and healthy controls.

## Methods

### Study cohorts and phenotypes

#### FinnGen

FinnGen is a large-scale biobank study that aims to genotype 500,000 Finnish participants recruited from hospitals as well as prospective and retrospective epidemiological and disease-based cohorts. These data are combined with longitudinal registries that record phenotypes and health events (including ICD-based diagnosis) over the entire lifespan including the National Hospital Discharge Registry (inpatient and outpatient), Causes of Death Registry, the National Infectious Diseases Registry, Cancer Registry, Primary Health Care Registry (outpatient) and Medication Reimbursement Registry. This study used data from FinnGen Data Freeze 7, which consisted of 309,154 individuals.

From the FinnGen population we identified patients with a CHD diagnosis and from those excluded individuals with a diagnosis for syndromes that are known to include CHD phenotypes. For the general CHD category, we used 2436 cases and 306 718 controls. In addition, the following CHD subgroups were analyzed: septal defects (atrial septal defects (ASD) and ventricular septal defects (VSD), N=1370), conotruncal defects (N=307) LVOTO narrow (LVOTO type congenital heart disease and coarctation of the aorta N=939), early aortic valve disease (including patients under 60 years of age with non-congenital aortic valve stenosis and/or insufficiency diagnosis, N=2183), and LVOTO broad (including individuals included in the LVOTO narrow and early aortic valve disease groups N=2313). The diagnoses used for each subgroup and the diagnoses used for exclusion are listed in Supplemental Table 1.

### FinnGen ethics statement

Patients and control subjects in FinnGen provided informed consent for biobank research, based on the Finnish Biobank Act. Alternatively, separate research cohorts, collected before the Finnish Biobank Act came into effect (in September 2013) and before start of FinnGen (August 2017), were collected based on study-specific consents and later transferred to the Finnish biobanks after approval by Fimea (Finnish Medicines Agency), the National Supervisory Authority for Welfare and Health. Recruitment protocols followed the biobank protocols approved by Fimea. The Coordinating Ethics Committee of the Hospital District of Helsinki and Uusimaa (HUS) statement number for the FinnGen study is Nr HUS/990/2017.

The FinnGen study is approved by the Finnish Institute for Health and Welfare (permit numbers: THL/2031/6.02.00/2017, THL/1101/5.05.00/2017, THL/341/6.02.00/2018, THL/2222/6.02.00/2018, THL/283/6.02.00/2019, THL/1721/5.05.00/2019 and THL/1524/5.05.00/2020), Digital and population data service agency (permit numbers: VRK43431/2017-3, VRK/6909/2018-3, VRK/4415/2019-3), the Social Insurance Institution (permit numbers: KELA 58/522/2017, KELA 131/522/2018, KELA 70/522/2019, KELA 98/522/2019, KELA 134/522/2019, KELA 138/522/2019, KELA 2/522/2020, KELA 16/522/2020), Findata permit numbers THL/2364/14.02/2020, THL/4055/14.06.00/2020,,THL/3433/14.06.00/2020, THL/4432/14.06/2020, THL/5189/14.06/2020, THL/5894/14.06.00/2020, THL/6619/14.06.00/2020, THL/209/14.06.00/2021, THL/688/14.06.00/2021, THL/1284/14.06.00/2021, THL/1965/14.06.00/2021, THL/5546/14.02.00/2020 and Statistics Finland (permit numbers: TK-53-1041-17 and TK/143/07.03.00/2020 (earlier TK-53-90-20)).

The Biobank Access Decisions for FinnGen samples and data utilized in FinnGen Data Freeze 7 include: THL Biobank BB2017_55, BB2017_111, BB2018_19, BB_2018_34, BB_2018_67, BB2018_71, BB2019_7, BB2019_8, BB2019_26, BB2020_1, Finnish Red Cross Blood Service Biobank 7.12.2017, Helsinki Biobank HUS/359/2017, Auria Biobank AB17-5154 and amendment #1 (August 17 2020), Biobank Borealis of Northern Finland_2017_1013, Biobank of Eastern Finland 1186/2018 and amendment 22 § /2020, Finnish Clinical Biobank Tampere MH0004 and amendments (21.02.2020 & 06.10.2020), Central Finland Biobank 1-2017, and Terveystalo Biobank STB 2018001.

### SAIGE GWAS analysis

The GWAS for the CHD categories was performed using SAIGE on the FinnGen R7 cohort (N=309 154) (Zhou et al., 2020). The primary settings for the SAIGE analysis were; ratio coefficient of variation (CV) cutoff: 0.001, trace CV cutoff: 0.0025, minmac: 5. The covariates used for the analysis included: sex, imputed age, age at death or end of follow up, 10 principal components and microarray genotyping batch (full list in Supplemental Table 2). The FinnGen variant filtering for PCA and GWAS analysis is: exclusion of variants with info score < 0.95, exclusion of variants with missingness > 0.01 (based on the GP; see conversion), exclusion of variants with MAF < 0.05 and LD pruning with window interval of 500kb and 50kb steps, and an r^2 filter of 0.1 (https://finngen.gitbook.io/documentation/methods/phewas/quality-checks).

### Sensitivity fine-mapping

GWAS significant variants were re-tested for association using REGENIE (Mbatchou et al., 2021) fine-mapped using SuSiE and FINEMAP (Benner et al., 2016; Wang et al., 2020). The general FinnGen fine-mapping pipeline is described (https://github.com/FINNGEN/finemapping-pipeline).

### Calculating eQTL data

To test whether the genome-wide significant SNPs from the FinnGen GWAS could have an effect on the expression of nearby genes, we utilized the eQTL calculator function of the GTEx portal database (www.gtexportal.org) (Lonsdale et al., 2013; Stanfill and Cao, 2021). We examined expression in the following tissues: ‘heart - atrial appendage’ and ‘heart – left ventricle’.

### Defining endpoint associations using PheWeb

In addition to CHD, FinnGen contains diagnosis codes and precomputed phenotypes. Therefore, instead of looking at each disease or GWAS alone, it is possible to examine a single significant variant horizontally across other diseases to understand which other diseases are associated with this variant. The FinnGen results website uses the PheWeb design interface for displaying SNP-endpoint associations for a convenient overview of GWAS results (Gagliano Taliun et al., 2020). This approach was used to identify other endpoints associated with the lead SNPs. The FinnGen R7 data will become available to the public similarly as release 5.

### Generation of pluripotent stem cell lines

Human induced pluripotent stem cells (hiPSCs) were reprogrammed at the Biomedicum Stem Cell Center Core Facility (University of Helsinki). The cell lines were derived from four male individuals with LVOTO defects, (three patients with HLHS and one patient with congenital aortic stenosis (AS) and coarctation of the aorta (COA)). The cell lines were created by using retroviral/Sendai virus transduction of Oct3/4, Sox2, Klf4, and c-Myc, as described previously (Trokovic et al., 2015). Three control cell lines were obtained from The Biomedicum Stem Cell Center Core Facility (University of Helsinki) and one control line was a kind gift from Professor Anu Suomalainen-Wartiovaara. All control cell lines were male. The cell lines were differentiated to hiPS-CMs as previously described (Helle et al., 2021). Briefly, Wnt-pathway activation and inactivation were used to induce differentiation, and metabolic selection with media containing lactate instead of glucose was used to ensure purity of the CM culture. The cells were processed for single cell sequencing as described previously (Helle et al., 2021). The cells were detached using Multi Tissue Dissociation Kit 3 (130-110-204, Miltenyi Biotec), according to the manufacturer’s instructions. The cells were washed once with PBS containing 0.04% BSA, and then resuspended in PBS with 0.04% BSA to a concentration of 2.0 × 106 cells/ml. The cells were passed through a 35 μm strainer (352235, Corning) and combined into two samples (one containing cells from healthy individuals, one containing patient cells) and placed on ice until continuation of the 10× Genomics Single Cell Protocol at the Institute of Molecular Medicine Finland (FIMM).

### Ethics statement of the hiPSC work

The hiPSC-study protocol was reviewed and approved by Coordinating Ethics Committee of the HUS Hospital District. Written informed consent was obtained from the legal guardians of the participants and also directly from all participants aged over 6 years.

### Single-cell RNA sequencing and analysis

Single-cell gene expression profiles were studied using the 10X Genomics Chromium Single Cell 3′RNAseq platform. The Chromium Single Cell 3′RNAseq run and library preparation were made using the Chromium Single Cell 3′ Reagent version 3 chemistry. The sample libraries were sequenced on Illumina NovaSeq 6000 system. 12000 cells/combined sample and 50,000 PE/cell were analyzed. Data processing and analysis were performed using 10X Genomics Cell Ranger v2.1.1 pipelines. The “cellranger mkfastq” pipeline was used to produce FASTQ (raw data) files. The “cellranger count” was used to perform alignment, filtering and UMI counting. Mkfastq was run using the Illumina bcl2fastq v2.2.0 and alignment was done against human genome GRCh38. Cellranger “aggr” pipeline was used to combine data from multiple samples into an experiment-wide gene-barcode matrix and analysis. The scRNAseq data was analyzed using the R package Seurat v3 (Satija et al., 2015). The data was filtered using a mitochondrial RNA percentage < 30, over 200 gene features per cell and at least 1000 identified RNA transcripts per cell. The control samples were pooled and used to compare against individual patients, and against the pooled patient data.

### HLA allele fine-mapping association with early aortic valve disease

We used the FinnGen imputed dosages of HLA alleles (HLA-A, B, C, DPB1, DQA1, DQB1, DRB1, DRB3, DRB4 and DRB5) for the R7 cohort. These alleles have been previously imputed using a Finnish specific imputation panel (Ritari et al., 2020). We then performed logistic regression using the binominal R generalized linear model (glm) function. As covariates, the principal components (PC) 1-20 for the cohort were used, and age, sex, age^2^ and age*sex.

## Results

### FinnGen GWAS highlights SNPs significantly associated with CHD

From the general CHD category GWAS we detected one lead SNP in chromosome 17, rs11570508 (beta = 0.24, P = 1.2×10^−11^, intronic region of CDC27) (Figure 1A,B, Table 1), and fine-mapping of the signal suggested this variant to be the primary causal variant (Supplemental Table 3). Furthermore, we detected GWAS significant lead SNPs on chromosome 17 associated with septal defects (rs1293973611, beta = 4.96, P = 1.1×10^−8^, intronic region of RP11-1055B8.3), chromosome 20 associated with LVOTO broad (rs1342740627, beta = 4.8, P = 1.8×10^−8^, intronic region of *GGTLC1*) and two SNPs in chromosomes 6 and 20 associated with early aortic valve disease (rs17214148, beta = 3.2, P = 3.4×10^−8^, 3 prime UTR-region of *HLA-DOA*; and rs1342740627, beta = 4.9, P = 3.4×10^−8^) (Supplemental Figure 2, Table 1). Sensitivity analysis using fine-mapping confirmed that the lead SNPs rs1293973611 SNP associated with septal defects, and the rs1342740627 SNP associated with LVOTO broad. These two SNPs were also found to be greatly enriched in the Finnish population (Table 1). GWAS diagnostics were assayed with QQ-plots (Supplemental Figure 3). No GWAS significant SNPs were identified for LVOTO narrow or conotruncal defects. To fine map the association with the *HLA-DOA* region, we performed a glm regression showed a significant (Bonferroni corrected for multiple testing) association for 6 HLA alleles and early aortic valve disease (Supplemental Table 4). The most significant was DRB1*01:02 (beta = 1.14, P = 6.9×10^−6^).

**Table 1.** GWAS significant lead SNPs found in FinnGen release 7 for several CHD groups.

| Subgroup | Variant | Location<br>(hg38) | Finnish<br>Enrichment | AF<br>cases | AF<br>controls | beta | SE | P-value |
| --- | --- | --- | --- | --- | --- | --- | --- | --- |
| CHD general | rs11570508 | 17:47151194 | 1.190 | 0.29 | 0.24 | 0.24 | 0.035 | $1.2 \times 10^{-11}$ |
| LVOTO broad | rs1342740627 | 20:23988232 | 1000000.000 | 0.0030 | 0.00054 | 4.79 | 0.85 | $1.78 \times 10^{-8}$ |
| Early aortic<br>valve disease | rs17214148 | 6:33004385 | 0.0770 | 0.0051 | 0.0013 | 3.23 | 0.59 | $3.38 \times 10^{-8}$ |
| Early aortic<br>valve disease | rs1342740627 | 20:23988232 | 1000000.000 | 0.0037 | 0.00074 | 4.92 | 0.89 | $3.44 \times 10^{-8}$ |
| Septal defects | rs1293973611 | 17:81382139 | 1000000.000 | 0.0061 | 0.00087 | 4.96 | 0.87 | $1.1 \times 10^{-8}$ |

**Figure 1.**
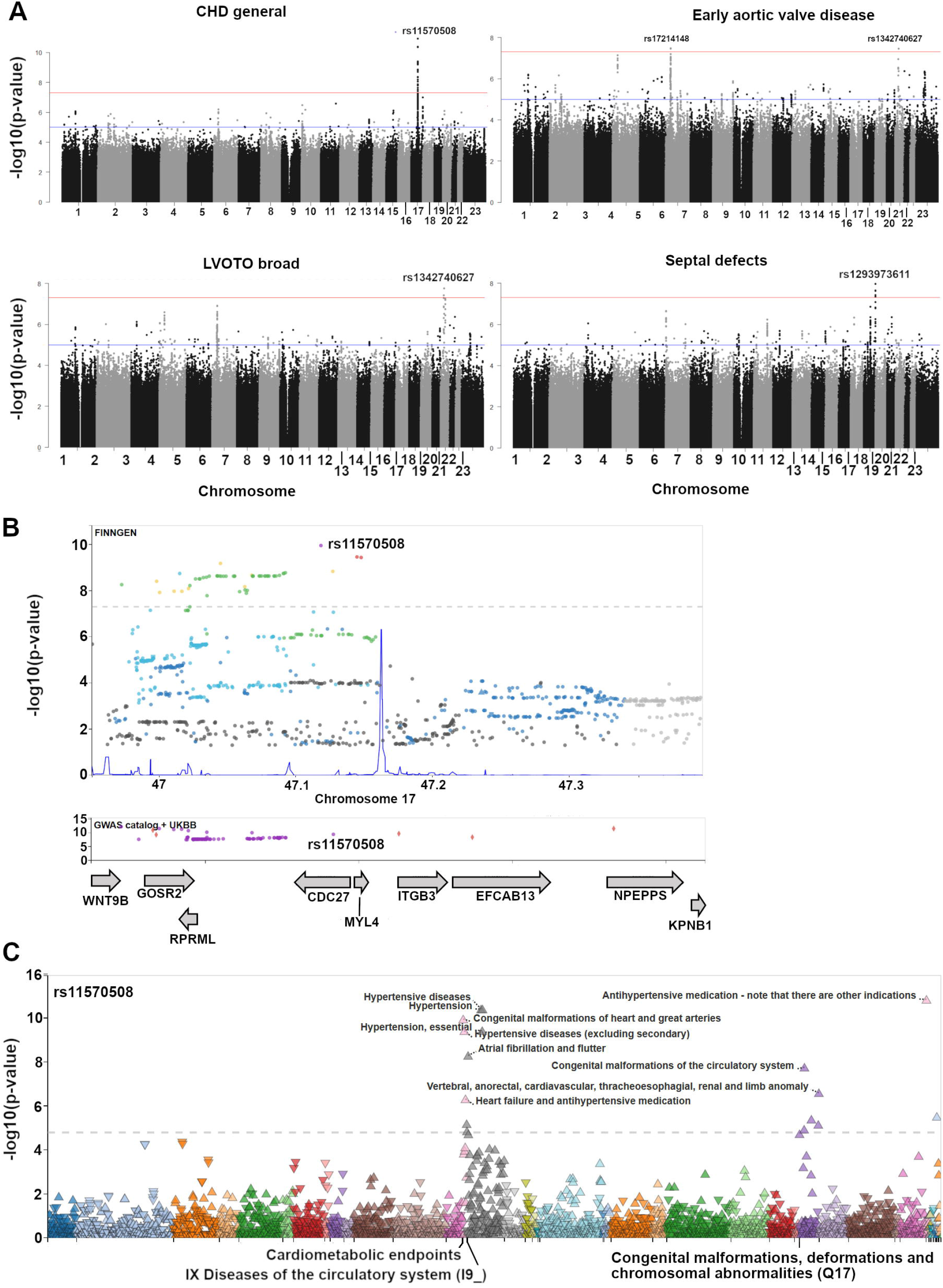
FinnGen R7 GWAS for general congenital heart disease (congenital malformations of the heart and great arteries) highlights rs11570508 as significantly associated. A; Genome-wide manhattan plots of CHD general, early aortic valve disease, LVOTO broad and septal defects with lead SNPs highlighted. B; Locus zoom plot with rs11570508 and nearby SNPs and genes. C; FinnGen PheWeb plot demonstrating rs11570508 associations with heart-based endpoints.

### eQTL analysis on the lead SNPs

The eQTL calculation demonstrated a potential effect by rs11570508 on the expression of two nearby genes: *MYL4* (eQTL P=0.0017 in heart - atrial appendage) and *KPNB1* (eQTL P=0.04 in heart - left ventricle). The eQTL calculation did not indicate significant effects on nearby genes for the other four GWAS significant SNPs. Thus, we decided to further explore the expression of *KPNB1* and *MYL4* in different scRNAseq datasets. Furthermore, a FUMA-based analysis predicted chromatin interactions using FUMA and associated eQTL data (Watanabe et al., 2017), suggesting an interaction between rs11570508 and *KPNB1* (Supplemental Figure 3).

### Genetic association results of FinnGen endpoints for the lead SNPs

In Figure 1C we show that rs11570508 is also significantly associated with other FinnGen cardiovascular endpoints, such as hypertensive diseases, atrial fibrillation and flutter, and use of antihypertensive medications (Figure 1C). The lead SNP for early aortic valve disease (rs17214148), is also significantly associated with the FinnGen endpoints for hypothyroidism disorders and disorders of the thyroid gland (Supplemental Figure 4). The other three lead SNPs did not have additional significant associations beyond CHD.

### Expression of *MYL4* and *KPNB1* in the cardiogenic region of early mouse embryos and in organotypic hiPSCs

Since our eQTL results suggested that rs11570508 (located in *CDC27*) could influence the expression of *MYL4* and *KPNB1* in the heart, we explored the expression of these two genes in published single-cell RNA seq data from the cardiogenic region of early mouse embryos (de Soysa et al., 2019). The data were visualized at the UCSC cell browser (Speir et al., 2021)(Figure 2A). The *KPNB1* homolog *Kpnb1* was expressed in all cells of the region, whereas the *MYL4* homolog *Myl4* was mainly expressed in the myocardium. Furthermore, we analyzed the expression of *MYL4* and *KPNB1* in hiPSC-derived endothelial cells (ECs) and cardiomyocytes (CMs) from a healthy individual grown in monoculture or in a coculture containing both cell types (Helle et al., 2021)(Figure 2B). The hiPS-ECs express the EC marker *KDR/VEGFR2*, and the hiPS-CMs express the cardiomyocyte marker *TNNT2*, defining the different cell types in the scRNAseq data (Figure 2B). As shown in Figure 2B, *MYL4* was highly expressed in the hiPS-CMs, and low expression levels were also detected in cardiac like hiPS-ECs that had been cultured together with hiPS-CMs (EC-CM Coculture), while no expression was detected in hiPS-ECs cultured alone (EC Monoculture). *KPNB1* was expressed in both cell types at similar levels, similar to what was observed in the embryonic mouse heart (Figure 2B).

**Figure 2.**
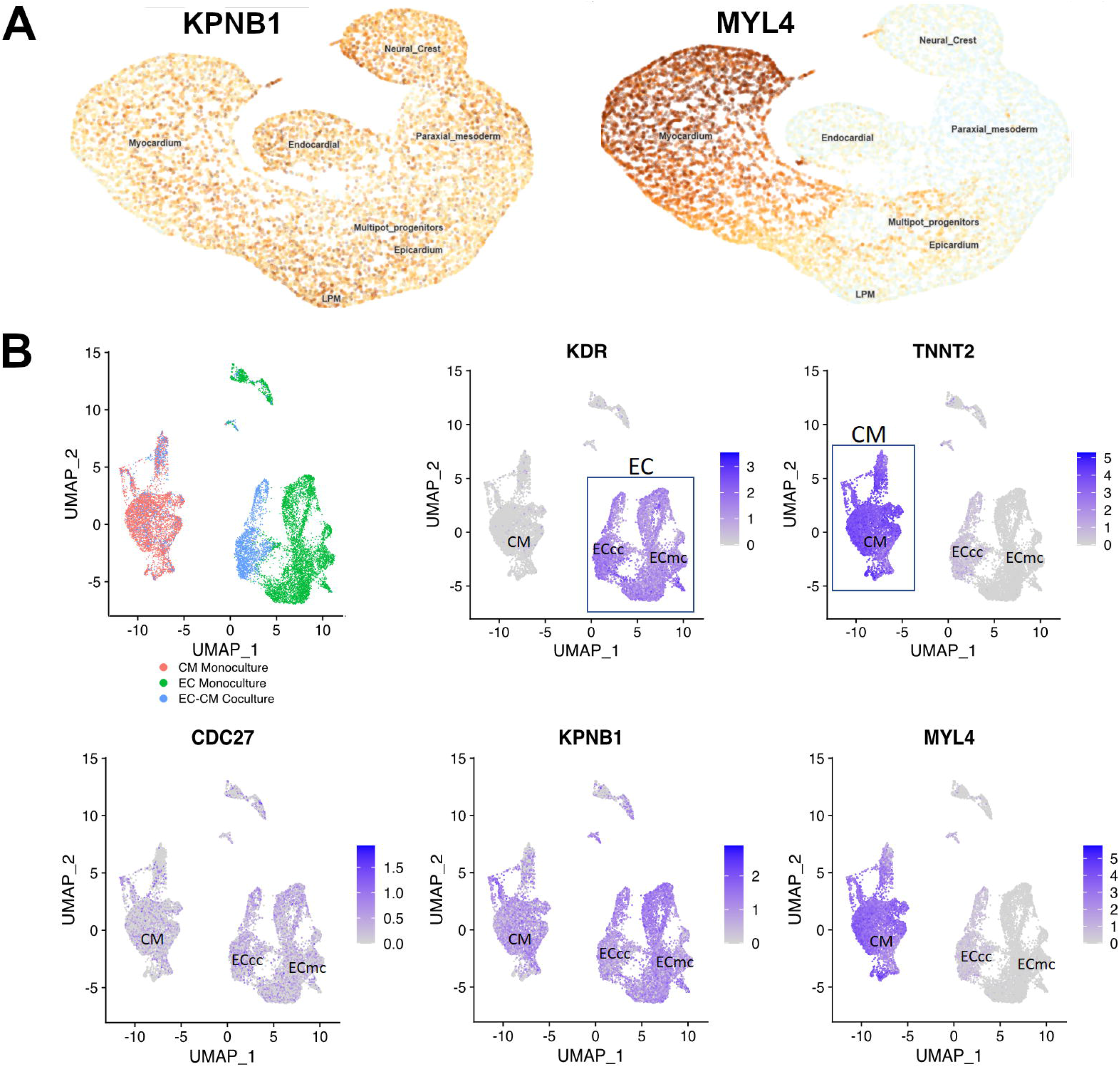
MYL4 and KPNB1 are expressed differently in cardiac cell types in mice and humans. A; early embryonic mouse cardiogenic map of KPNB1 and MYL4 cell expression based on the study by de Soysa et al. (2019), as visualized on the UCSC cell browser website. B; scRNAseq gene expression UMAP plots from cardiomyocyte-endothelial cell cultures (Helle etal. 2019) for KDR, TNNT2, MYL4, KNPB1 and CDC27.

### HiPS-CMs derived from CHD patients have lower expression of *MYL4* and *KPNB1*

Next, we compared the expression of *MYL4* and *KPNB1* in scRNAseq data from hiPS-CMs differentiated from hiPSCs of four LVOTO patients and four healthy controls. We observed that the expression levels of *KPNB1* and *MYL4* were lower in three of the four LVOTO patients as compared with healthy controls (Figure 3B). Comparing the pooled patient samples to controls also showed a significant downregulation of KPNB1 as well: log2 fold change (log2FC) = -0.4, P = 5.2×10^.49^.

**Figure 3.**
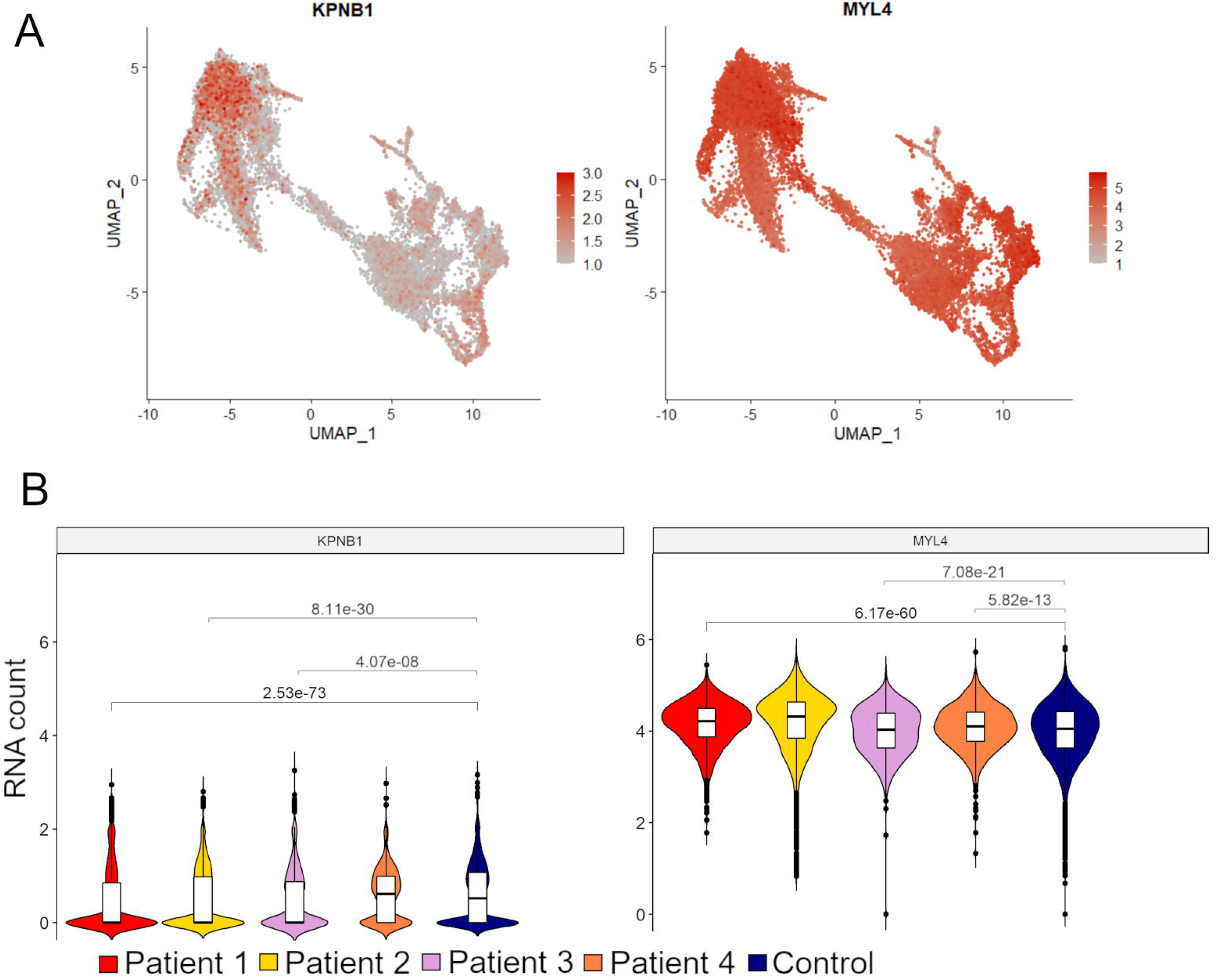
KPNB1 and MYL4 are significantly down-regulated LVOTO patients’ hiPSC-CMs compared to healthy controls. A; UMAP plot of the KPNB1 and MYL4 expression across all cells. B; violin plot and box plots displaying the scaled RNA counts of KPNB1 and MYL4 in all 4 patient samples and the control pool sample.

## Discussion

Here we report the results of a GWAS of general CHD traits in the FinnGen R7 database, as well as of the CHD subgroups; septal defects, LVOTO (broad group) and early aortic valve disease (age < 60 years). We detected one genome-wide significant SNP associated with CHD in general, and four genome-wide significant SNPs for the subgroups. According to eQTL analysis, the general CHD associated SNP rs11570508 associated with the expression of *KPNB1* and *MYL4* in heart tissue, and a chromatin interactions analysis suggested an interaction between rs11570508 and *KPNB1*. Further, we demonstrated that the expression of these genes was lower in CHD patient-derived iPSC-cardiomyocytes compared to healthy controls.

Two SNPs, rs1293973611 associated with septal defects, and rs1342740627 associated with LVOTO broad and early aortic valve disease, were highly enriched in the Finnish population. These loci have not been associated with CHD or aortic valve disease previously. The finding highlights the value of the FinnGen project in identifying novel rare or low-frequency disease variants in an isolated population with enrichment of variants due to recent population bottlenecks followed by rapid population growth (Table 1).

The SNP rs11570508 associated with general CHD is located in a region that contains several genes linked to heart development, CHD and other cardiac phenotypes; *MYL4, CDC27, GOSR2* and *WNT9B* (Goddard et al., 2017; Lahm et al., 2021; Orr et al., 2015; Sabour et al., 2018). A recent GWAS study on patients with anomalies of thoracic arteries and veins identified three significant SNPs within the *GOSR2* locus in chromosome 17 close to rs11570508 (Lahm et al., 2021), and the chr17:45013271_T_C (rs17608766) variant in *GOSR2* has been associated with a reduced aortic valve area (Córdova-Palomera et al., 2020) indicating importance of this locus in cardiac development.

In the eQTL database GTEx (Analysis Release V8 dbGaP Accession phs000424.v8.p2), rs11570508 was predicted to influence the expression of *KPNB1* and *MYL4* in the heart. *KPNB1* encodes nuclear import receptor karyopherin β1, which is a nuclear importer protein involved in the translocation of proteins and part of the regulation of mitosis (Zhu et al., 2018). It has not been linked to CHD previously, but has been reported as a potential oncogene (Kodama et al., 2017). Our analysis showed that *KPNB1* homolog is expressed in the developing cardiogenic region in the developing murine heart, and hiPSC-derived CMs and ECs, suggesting that it might have a role during embryonic development in CHD-relevant cell types.

*MYL4* encodes the atrial light chain-1 (ALC-1) protein, which is expressed in the human heart during development (Barton and Buckingham, 1985). After birth, the expression decreases in the ventricles but remains in the atria (Wang et al., 2018). ALC-1 has a role in sarcomere assembly and fine tuning of cardiac contractility. The expression of MYL4 has shown to abnormally persist in ventricular tissue of individuals with CHD (England and Loughna, 2013), and the ventricular analog *MYL3* is replaced by re-expression of *MYL4* in failing and hypertrophied hearts, resembling a fetal remodeling pattern associated with ventricular dysfunction (Wang et al., 2019, 2018). *MYL4* loss-of-function variants have been associated with atrial cardiomyopathy and atrial arrhythmias such as atrial fibrillation (AF)(Gudbjartsson et al., 2017, 2015; Orr et al., 2015; Peng et al., 2017), demonstrating that although previously not associated with CHD directly, its role in cardiac development and function is important, and thus, could be a plausible candidate gene for CHD.

Lower expression levels of *MYL4* and *KPNB1* were seen in the scRNAseq data of three of four hiPS-CM cell lines derived from LVOTO patients as compared to heathy controls. Recent studies have indicated that despite the genetic heterogeneity, similar abnormal phenotypes and gene expression profiles in LVOTO patient derived hiPS-CMs can be observed, indicating for example diminished capacity for cardiomyocyte differentiation, more immature transcriptomic profile, and reduced expression of cardiac transcription factors (Jiang et al., 2014; Kobayashi et al., 2014; Krane et al., 2021; Theis et al., 2015; Yang et al., 2017). The lower expression levels of *MYL4* and *KPNB1* in patient-derived cells may, thus, indicate a role of these genes in the development of CHD.

Interestingly, in addition to CHD, the FinnGen Atrial fibrillation (AF) and flutter, and Hypertensive diseases endpoints were significantly associated with chr17:47151194:C_A (rs11570508). Children and adults with CHD have an increased risk for AF (Labombarda et al., 2017; Mandalenakis et al., 2018), and adults with CHD have increased cardiovascular morbidity relative to the general population (Saha et al., 2019). Although anatomical factors such as anomalous vessel anatomy and abnormal atrial hemodynamics, and disease related conditions in the heart such as progressive valvulopathy, residual shunts, and atrial scars from previous heart surgery are likely to be important predisposes for AF and other cardiovascular morbidity in CHD, it is tempting to speculate that shared genetic factors could potentially increase the risk for both developing CHD and later cardiovascular morbidity. In fact, atrial arrythmias are a common phenotype in Holt-Oram syndrome, a developmental disorder that leads to heart and limb malformations. Holt-Oram syndrome is partially caused by variants in the *TBX5* gene (Basson et al., 1997; Vanlerberghe et al., 2019), and GWAS studies on AF have shown several significant loci with nearest genes known to be important heart developmental and causal for CHD, such as *NKX2-5, GATA4, GJA1, and GJA5* (Pierpont et al., 2018; Roselli et al., 2020). It has been also shown that women whose infants have congenital heart defects have an increased risk and earlier disease onset for cardiovascular morbidity, including atherosclerotic cardiovascular disease, ischemic heart disease, hospitalization due to cardiac diseases and cardiac transplantation (Auger et al., 2018). Thus, these associations could indicate shared genetic risks.

The eQTL analysis did not identify associations for genes expressed in adult cardiac tissues for the rs1293973611 variant associated with septal defects, the rs1342740627 variant associated with LVOTO broad, or the rs17214148 or rs1342740627 variants associated with early aortic valve disease. In addition, their nearest genes have not been associated with CHD before. Thus, at present, we do not have evidence for the functional causality for these variants. However, it is well known that the gene expression in the adult heart is significantly different from that of the development of cardiac structures, and thus absence of association with adult heart gene expression does not rule out potential association with gene expression in the embryonic heart. Interestingly, the beta values for all these three very rare variants were quite big, potentially indicating large effect sizes. This finding has important implications for the genetic architecture of CHD suggesting that rare variants can be associated with a substantial risk for rare disease such as CHD.

Bicuspid aortic valve disease, and other less severe developmental aortic valve defects can be asymptomatic during childhood and cause aortic valve insufficiency or stenosis later in life. Calcific aortic valve stenosis manifests typically during the eight decade of life (Martinsson et al., 2015), and it is likely that a developmental defect underlies cases diagnosed at younger ages suggesting two different disease entities. Thus, in addition to those with CHD diagnosis, we created a subgroup of those who received aortic valve stenosis or insufficiency diagnosis under the age of 60 years. GWAS loci in genes such as LPA and PALMD have been repeatedly associated with calcific aortic valve stenosis (Helgadottir et al., 2018; Thanassoulis et al., 2013; Thériault et al., 2018), however, those associations were not found in our study. Instead, two other significant loci were identified, of which one was significant even in the LVOTO broad group including patients with congenital LVOTO diagnoses. This finding could suggest a distinct genetic and pathophysiological etiology for early onset aortic valve disease in this population, which could be closer to that of CHD.

Our approach has some limitations. The discovery of novel loci in GWAS studies does not prove functional causality, and validation studies should be conducted. In addition, ICD10-based studies are limited by the fact that mild CHD such as BAV and hemodynamically insignificant ASD are often underdiagnosed as they may be asymptomatic. In addition, our association results are from the isolated Finnish population, which is genetically unique, and some of the identified variants were enriched in the Finnish population preventing replication in other populations. On the other hand, our study highlights the potential of FinnGen to identify disease associated high-impact variants that are very rare or absent in other populations.

In summary, our analysis of 2436 cases and 306718 controls identifies a robust risk locus for CHD near candidate genes with a known role in heart development, and *MYL4* and *KPNB1* were demonstrated as potentially causal genes for CHD. In addition, three risk loci for three CHD subtypes had a high beta value, suggesting that very rare variants can play a substantial role in CHD. The associations between the identified locus and other cardiovascular conditions, such as AF and flutter and hypertension suggest shared genetic etiology in developmental defects and adult cardiovascular morbidity.

## Data Availability

ScRNAseq data files used for gene expression analysis will be made available in a public repository prior to publication.

## Acknowledgements

This work was funded by grants from the Finnish Cultural Foundation (E.H, T.O. and M.B.), Finnish Foundation for Pediatric Research (E.H, T.O and M.B.), the Finnish Medical Foundation (E.H), Helsinki University (E.H), Helsinki University Hospital (E.H), and The Academy of Finland (E.H). Furthermore, we would like to thank the FinnGen consortium for GWAS summary statistics. We also thank the FIMM Single-Cell Analytics unit supported by HiLIFE and Biocenter Finland for single cell services.

## Supplemental Files

**Supplemental Document 1. Expanded FinnGen contributor list**.

**Supplemental Table 1. ICD-10 codes used as end-points for the main CHD categories in this study**.

**Supplemental Table 2. List of covariates used in the SAIGE GWAS analysis**.

**Supplemental Table 3. FinnGen SuSiE and FINEMAP results for the lead SNPs**.

**Supplemental Table 4. Most significant HLA alleles from the generalized linear model regression analysis against Early aortic valve disease**.

**Supplemental Figure 1. Locus zoom plots (v0.13.3) of the lead SNPs for LVOTO broad, aortic valve disease early age (<60 years) and septal defects**.

**Supplemental Figure 2. QQ plots from the GWAS analyses of the different CHD groups**.

**Supplemental Figure 3. Circos plot displaying a putative chromatin interaction (orange link) between KPNB1 and rs11570508**

**Supplemental Figure 4. FinnGen PheWeb plot of rs17214148 with most significantly associated end points highlighted**.

