## Supplemental Document 1 for "Genomic and transcriptomic data analyses highlight *KPNB1* and *MYL4* as novel risk genes for congenital heart disease"

### Acknowledgements for FinnGen data using manuscripts (by Mervi Aavikko 2.11.2021)

**Acknowledgements:**

We want to acknowledge the participants and investigators of FinnGen study. The FinnGen project is funded by two grants from Business Finland (HUS 4685/31/2016 and UH 4386/31/2016) and the following industry partners: AbbVie Inc., AstraZeneca UK Ltd, Biogen MA Inc., Bristol Myers Squibb (and Celgene Corporation & Celgene International II Sàrl), Genentech Inc., Merck Sharp & Dohme Corp, Pfizer Inc., GlaxoSmithKline Intellectual Property Development Ltd., Sanofi US Services Inc., Maze Therapeutics Inc., Janssen Biotech Inc, Novartis AG, and Boehringer Ingelheim. Following biobanks are acknowledged for delivering biobank samples to FinnGen: Auria Biobank (www.auria.fi/biopankki), THL Biobank (www.thl.fi/biobank), Helsinki Biobank (www.helsinginbiopankki.fi), Biobank Borealis of Northern Finland (<https://www.ppshp.fi/Tutkimus-ja-opetus/Biopankki/Pages/Biobank-Borealis-briefly-in-English.aspx>), Finnish Clinical Biobank Tampere (www.tays.fi/en-US/Research_and_development/Finnish_Clinical_Biobank_Tampere), Biobank of Eastern Finland (www.ita-suomenbiopankki.fi/en), Central Finland Biobank (www.ksshp.fi/fi-FI/Potilaalle/Biopankki), Finnish Red Cross Blood Service Biobank (www.veripalvelu.fi/verenluovutus/biopankkitoiminta) and Terveystalo Biobank ([www.terveystalo.com/fi/Yritystietoa/Terveystalo-Biopankki/Biopankki/](http://www.terveystalo.com/fi/Yritystietoa/Terveystalo-Biopankki/Biopankki/)). All Finnish Biobanks are members of BBMRI.fi infrastructure ([www.bbmri.fi](http://www.bbmri.fi/)). Finnish Biobank Cooperative -FINBB (<https://finbb.fi/>) is the coordinator of BBMRI-ERIC operations in Finland. The Finnish biobank data can be accessed through the Fingenious^®^ services (<https://site.fingenious.fi/en/>) managed by FINBB.
