## Supplementary figures and images for "Genomic and transcriptomic data analyses highlight *KPNB1* and *MYL4* as novel risk genes for congenital heart disease"

### Supplemental Figure 1

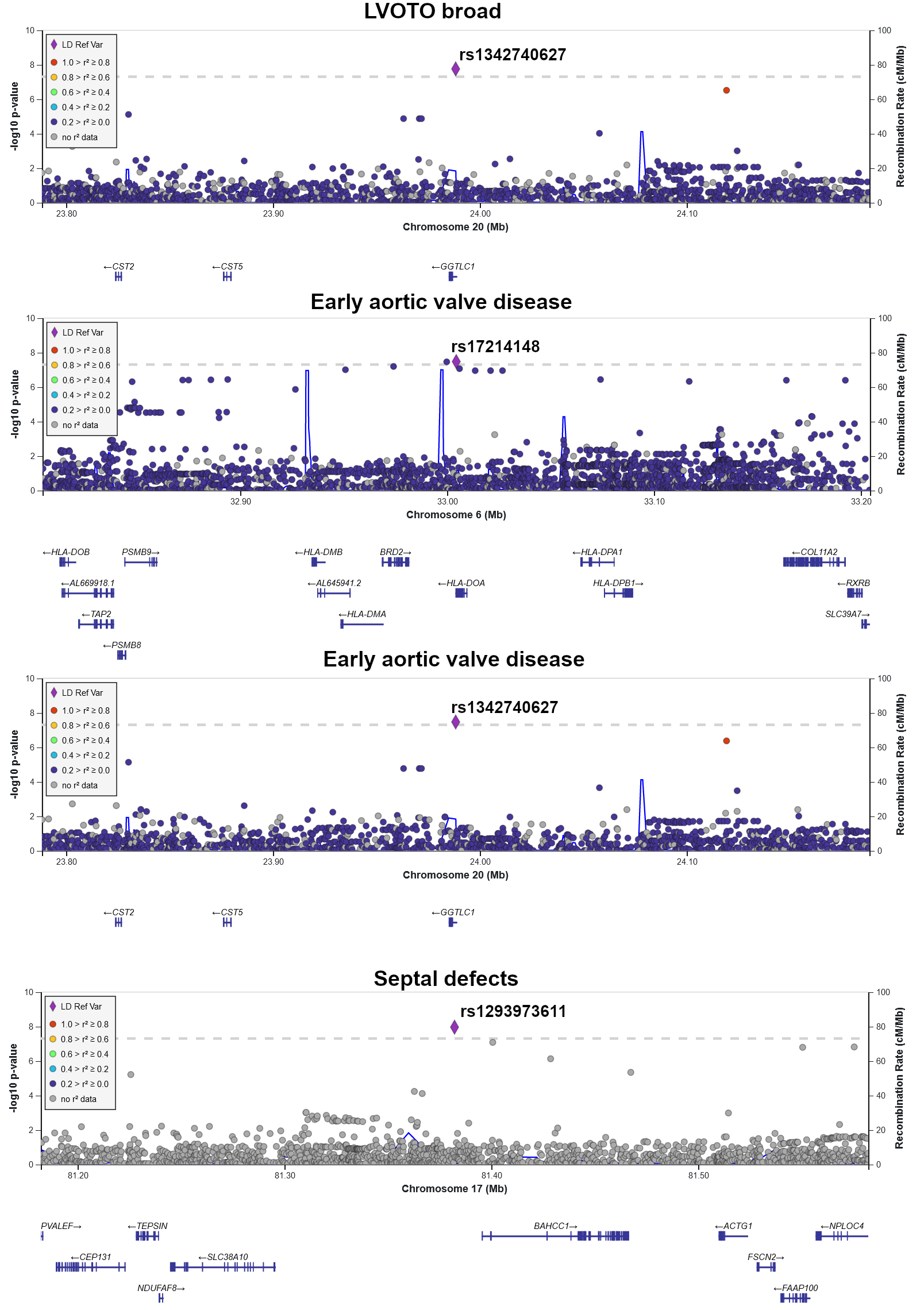

### Supplemental Figure 2

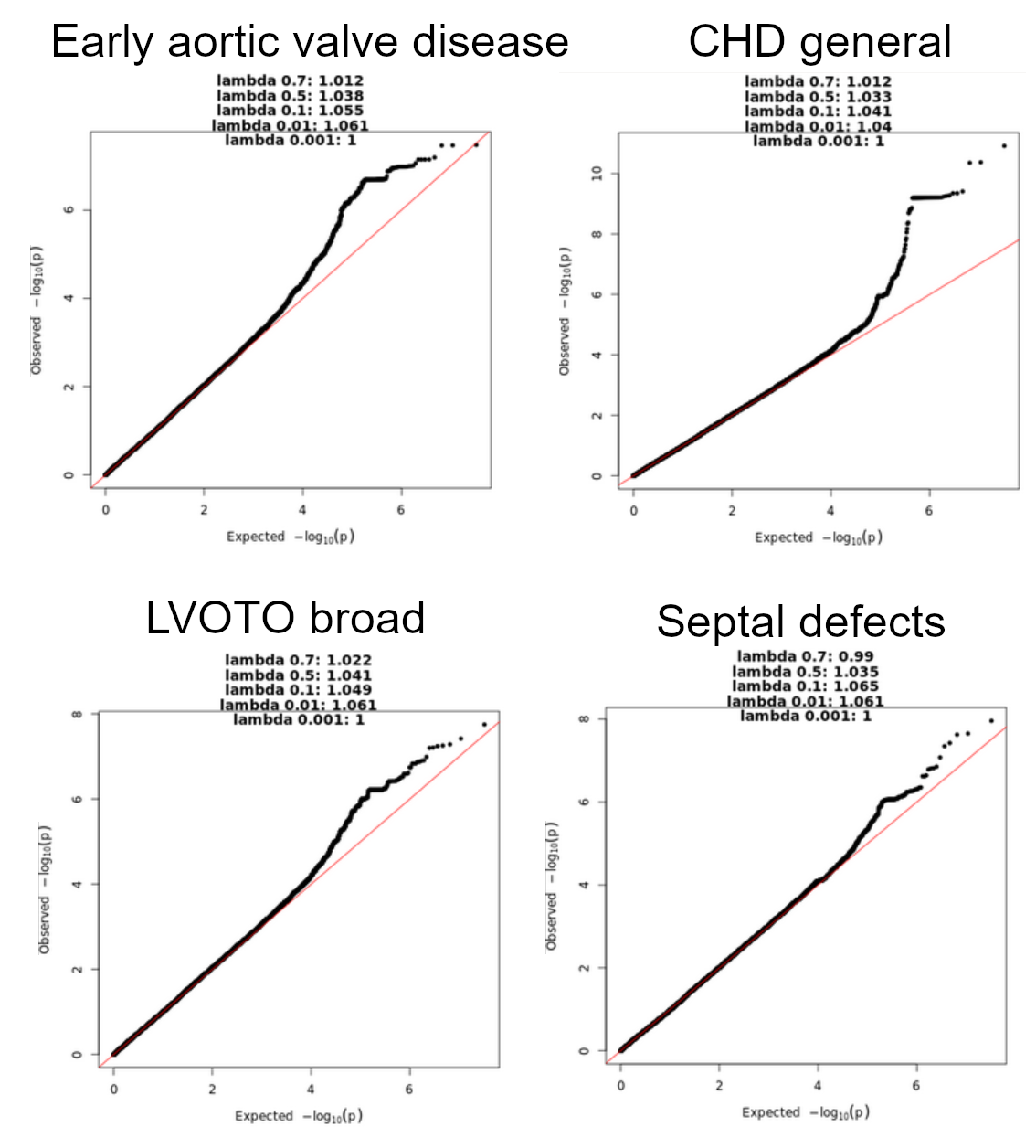

### Supplemental Figure 3

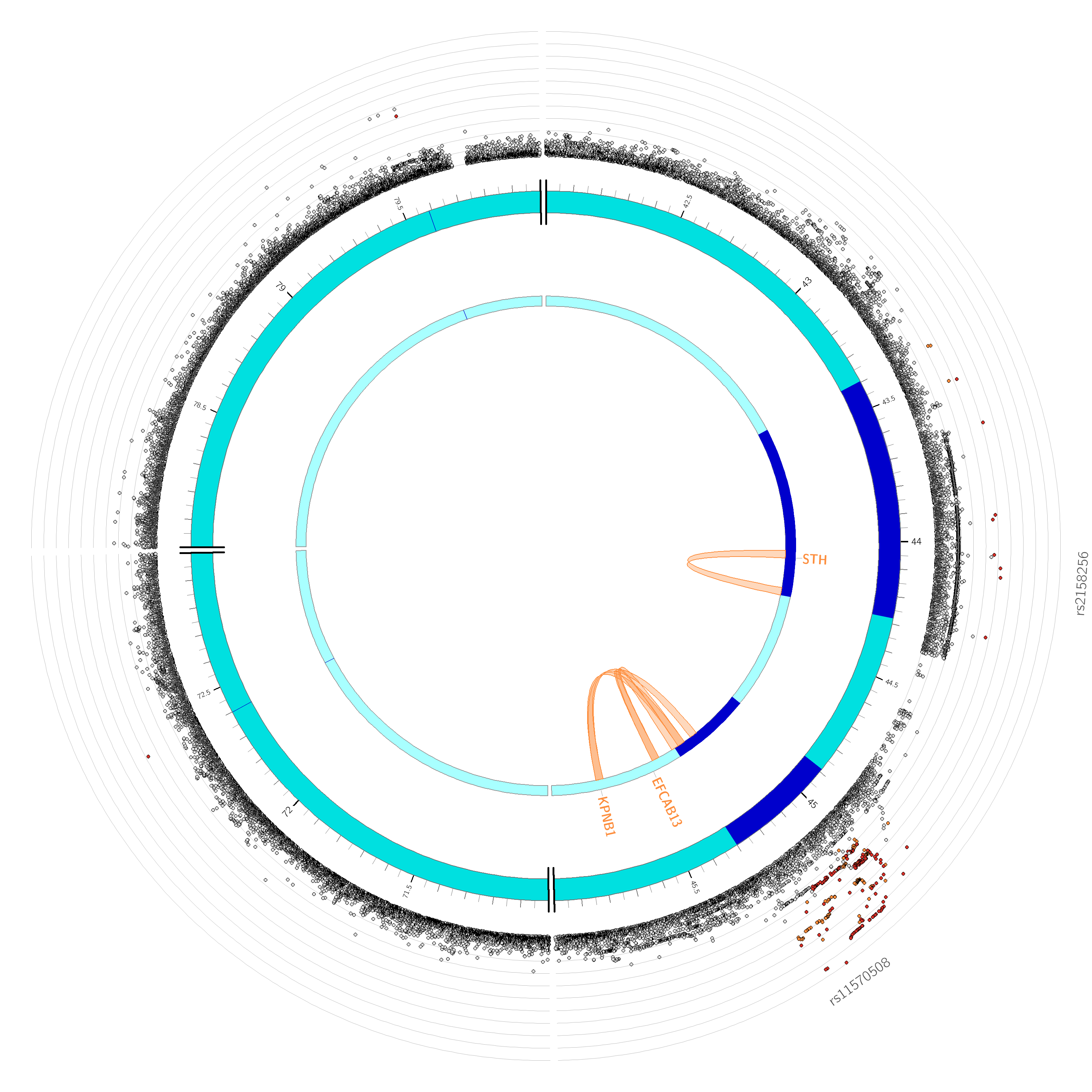

### Supplemental Figure 4

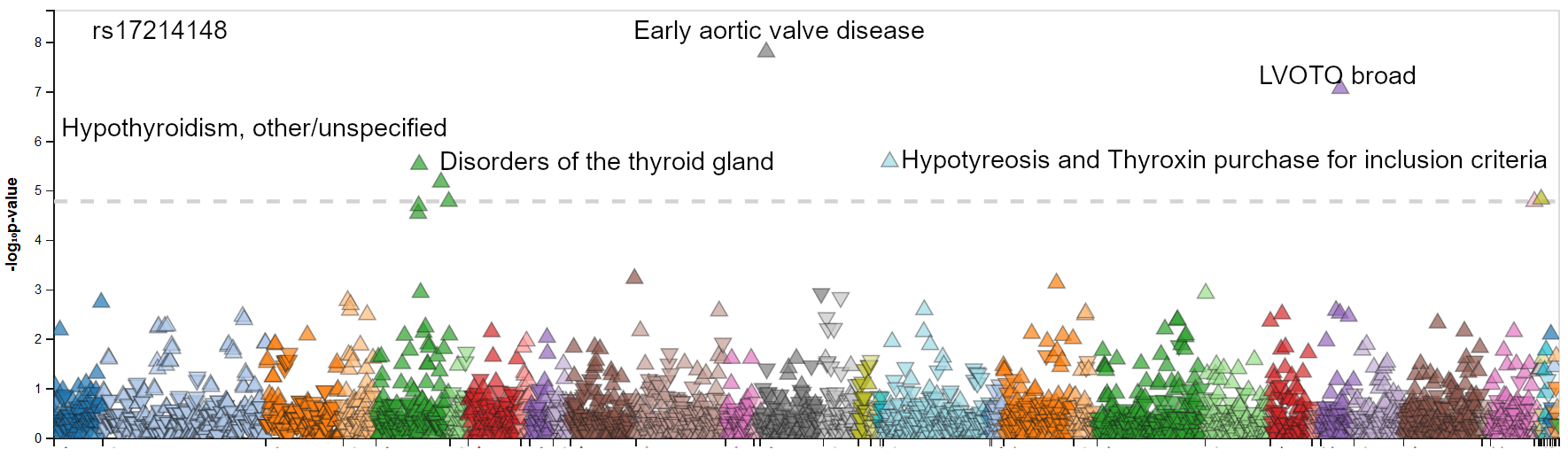
